# Microfluidic nano-scale qPCR enables ultra-sensitive detection of SARS-CoV-2

**DOI:** 10.1101/2020.08.28.20183970

**Authors:** Xin Xie, Tamara Gjorgjieva, Zaynoun Attieh, Mame Massar Dieng, Marc Arnoux, Mostafa Khair, Yasmine Moussa, Fatima Al Jallaf, Nabil Rahiman, Christopher A. Jackson, Zyrone Victoria, Mohammed Zafar, Raghib Ali, Fabio Piano, Kristin C. Gunsalus, Youssef Idaghdour

## Abstract

**Background:** A major challenge in controlling the COVID-19 pandemic is the high false-negative rate of the commonly used standard RT-PCR methods for SARS-CoV-2 detection in clinical samples. Accurate detection is particularly challenging in samples with low viral loads that are below the limit of detection (LoD) of standard one- or two-step RT-PCR methods.

**Methods:** We implement a three-step approach for SARS-CoV-2 detection and quantification that employs reverse transcription, targeted cDNA preamplification and nano-scale qPCR based on the Fluidigm 192.24 microfluidic chip. We validate the method using both positive controls and nasopharyngeal swab samples.

**Results:** Using SARS-CoV-2 synthetic RNA and plasmid controls, we demonstrate that the addition of a preamplification step enhances the LoD of the Fluidigm method by 1,000-fold, enabling detection below 1 copy/μl. We applied this method to analyze 182 clinical NP swab samples previously diagnosed using a standard RT-qPCR protocol (91 positive, 91 negative) and demonstrate reproducible detection of SARS-CoV-2 over five orders of magnitude (< 1 to 10^6^ viral copies/μl). Crucially, we detect SARS-CoV-2 with relatively low viral load estimates (<1 to 40 viral copies/μl) in 17 samples with negative clinical diagnosis, indicating a potential false negative rate of 18.7% by clinical diagnostic procedures.

**Conclusion:** The three-step nano-scale RT-qPCR method can robustly detect SARS-CoV-2 in samples with relatively low viral loads (< 1 viral copy/μl) and has the potential to reduce the false negative rate of standard RT-PCR-based diagnostic tests for SARS-CoV-2 and other viral infections.

**Summary:** We test, implement and report the results of a microfluidic RT-qPCR assay system involving sequential RT, preamplification and nano-scale qPCR that can robustly detect SARS-CoV-2 in clinical samples with viral loads less than 1 copy/ul.

## Introduction

The most widely used method for the detection of SARS-CoV-2 (the infectious agent that causes COVID-19) in nasopharyngeal (NP) swab samples is Reverse Transcription Polymerase Chain Reaction (RT-PCR) [1-3]. Inaccurate test results from RT-PCR have been widely reported, with estimated false negative rates of 10-30% among different implementations of this method [4-7]. Such high false negative rates pose a significant challenge to controlling the spread of infection worldwide, and are further exacerbated by poor sample quality or low viral loads that are below the detection limit of standard RT-PCR methods [8-9]. This, combined with both cost and scarcity of reagents [10-12] have hampered global scale-up of RT-PCR testing to levels that would be required to adequately monitor communities for COVID-19.

An additional challenge to controlling the spread of COVID-19 is the role of asymptomatic transmission [13-16]. Different estimates suggest that 40 to 80% of infected individuals are either pre-symptomatic, asymptomatic or mildly symptomatic [17-20]. Early detection of infection in these individuals is crucial for disease control, prompting many countries and communities to implement active screening programs that extend COVID-19 testing to asymptomatic individuals. However, asymptomatic carriers sometimes carry very low viral loads [21] that may not be detected by a standard RT-PCR test [22]. Therefore, the development of more sensitive detection methods that can detect low viral loads is crucial.

Most commercial kits for COVID-19 testing utilize either a one-step RT-PCR approach, which combines the RT and qPCR reactions, or a two-step approach in which RT and qPCR are performed sequentially. Here, we implement a three-step approach involving sequential RT, cDNA preamplification, and qPCR, using the Fluidigm microfluidics platform. Using this method, we demonstrate reliable ultra-sensitive detection of low SARS-CoV-2 viral loads in both standard positive controls and clinical NP swab samples, including samples previously diagnosed as negative by an accredited diagnostic lab. This microfluidic RT-PCR assay is a cost-effective strategy with the potential to reduce the false negative rate of clinical diagnostic tests, and as such, could be a valuable tool in active screening programs aiming at the early detection of SARS-CoV-2.

## Materials and Methods

We first implemented the three-step detection method using synthetic SARS-CoV-2 RNA and SARS-CoV-2 plasmids as standard positive controls. We then validated this method in clinical NP swab samples.

### Positive controls

Two types of positive controls were used in this study. The Twist Synthetic SARS-CoV-2 RNA (102024, Twist Biosciences) consists of six non-overlapping ssRNA fragments with the coverage of greater than 99.9% of the viral genome. The SARS-CoV-2 plasmids (10006625, IDT) contain the complete DNA sequence of the SARS-CoV-2 nucleocapsid gene.

### SARS-CoV-2 detection (Synthetic RNA and plasmid)

Two assays (primer/probe sets) were used for SARS-CoV-2 detection, per CDC recommendations: 2019-nCoV_N1 and 2019-nCoV_N2 (2019-nCoV CDC EUA Kit, 10006606, IDT). The human RNase P (RP) assay was used as a control for RNA extraction and RT-qPCR reactions. For both positive controls, 10-fold serial dilutions were prepared, with two replicates at each concentration. Each sample was analyzed using 9 replicates for N1, 9 replicates for N2, and 6 replicates for RP assays.

Manual extraction of synthetic SARS-CoV-2 RNA was performed using the ABIOpureTM Viral DNA/RNA Extraction Kit (M561VT50) according to the manufacturer’s instruction. The isolated synthetic RNA/SARS-CoV-2 plasmids were then used for reverse transcription (RT), 20 cycles of preamplification and quantitative PCR (qPCR) using the Fluidigm Real-Time PCR for Viral RNA Detection protocol (FLDM-00103, Fluidigm). qPCR mix of each sample and assays were loaded in the 192.24 integrated fluid circuit (IFC) chip. The chip was then placed in an IFC controller RX machine to pre-load the samples and the assays, and loaded onto the BioMark HD instrument (Fluidigm) for 35 cycles of qPCR quantification. A total of 4608 reactions were performed in each chip. The raw amplification data were acquired using the Fluidigm data collection software and analyzed using the Fluidigm Real-Time PCR Analysis software.

### Clinical samples and SARS-CoV-2 clinical diagnostics

A total of 182 de-identified nasopharyngeal (NP) swab samples (91 positive and 91 negative for SARS-CoV-2) were obtained from an accredited diagnostic lab. The diagnostic lab used the automated NX-48S Viral RNA Kit for RNA extraction, and the U-TOP^TM^ COVID-19 Detection Kit for SARS-CoV-2 detection, following accredited protocols in the United Arab Emirates that follow guidelines by the US Center for Disease Control and Prevention (CDC). Briefly, three primers were used, two targeting the viral ORF1ab and the N genes, and one acting as an internal control. A sample was classified as positive if at least one of the viral genes was detected with a Ct value ≤ 38. The reported LoD of the kit was 10 copies/reaction, translating to 1 copy/μl in the RNA sample.

### Three-step analysis of SARS-CoV-2 in clinical samples

Automated extraction of viral RNA from the clinical samples was performed using the Chemagic 360 automated nucleic acid extraction system and the Chemagic Viral DNA/RNA 300 Kit H96 (Perkin Elmer, CMG-1033S) according to the manufacturer’s instruction. 300μl clinical samples were used for RNA extraction and eluted in 80μl elution buffers. Subsequent RT, preamplification, and qPCR were performed as described above. Clinical samples were loaded onto two 96-well plates for RT and preamplification, with each plate containing 10-fold serially diluted SARS-CoV-2 plasmid controls (50-5000 copies/μl in the original experiment; 10-10,000 copies/μl in the replication experiments) used for viral load estimation. Samples were considered valid if the RP gene was reliably detected in at least 4 of the 6 replicates. Samples were classified as positive if at least one of the N assays (N1 or N2) was detected in at least one replicate. Each PCR plate also contained two negative controls: empty transport medium, to control for contamination during RNA extraction (NRX control), and TE dilution buffer, to control for contamination during pre-amplification and qPCR (NQF control).

### Statistical analysis

Data analysis was performed with R, associated packages, and GraphPad Prism 8. Virus copies were quantified based on a 10-fold dilution series of SARS-CoV-2 plasmids to generate standard curves. The standard curves were used to build log-linear models used to predict viral loads based on Ct values.

## Results

The objective of this study was to evaluate whether the microfluidic nano-scale RT-qPCR system has the potential to enhance the limit of detection (LoD) of SARS-CoV-2 in clinical samples. To do this, we first used synthetic SARS-CoV-2 RNA (Twist RNA) and SARS-CoV-2 plasmids to develop and evaluate our protocols (Figure 1A) and subsequently applied the method to 182 NP samples that were previously analyzed in a clinical diagnostic lab using standard RT-PCR protocols (Figure 1B). For all our experiments, we used the Fluidigm 192.24 microfluidic chip, which allows 192 samples to be independently analyzed against 24 different qPCR probes (24 assays), totaling 4,608 nano-scale qPCR reactions per run (Figure S1A). Per the US Center for Disease Control (CDC) standards, we used two qPCR probes targeting different regions of the N gene (N1 and N2) for SARS-CoV-2 detection (Figure S1B) and a probe targeting the human Ribonuclease P (RP) gene as a quality control. To increase the robustness of detection for each assay, we performed 9 technical replicates for the N1 and N2 assays, and 6 replicates for the RP assay.

**Figure 1.**
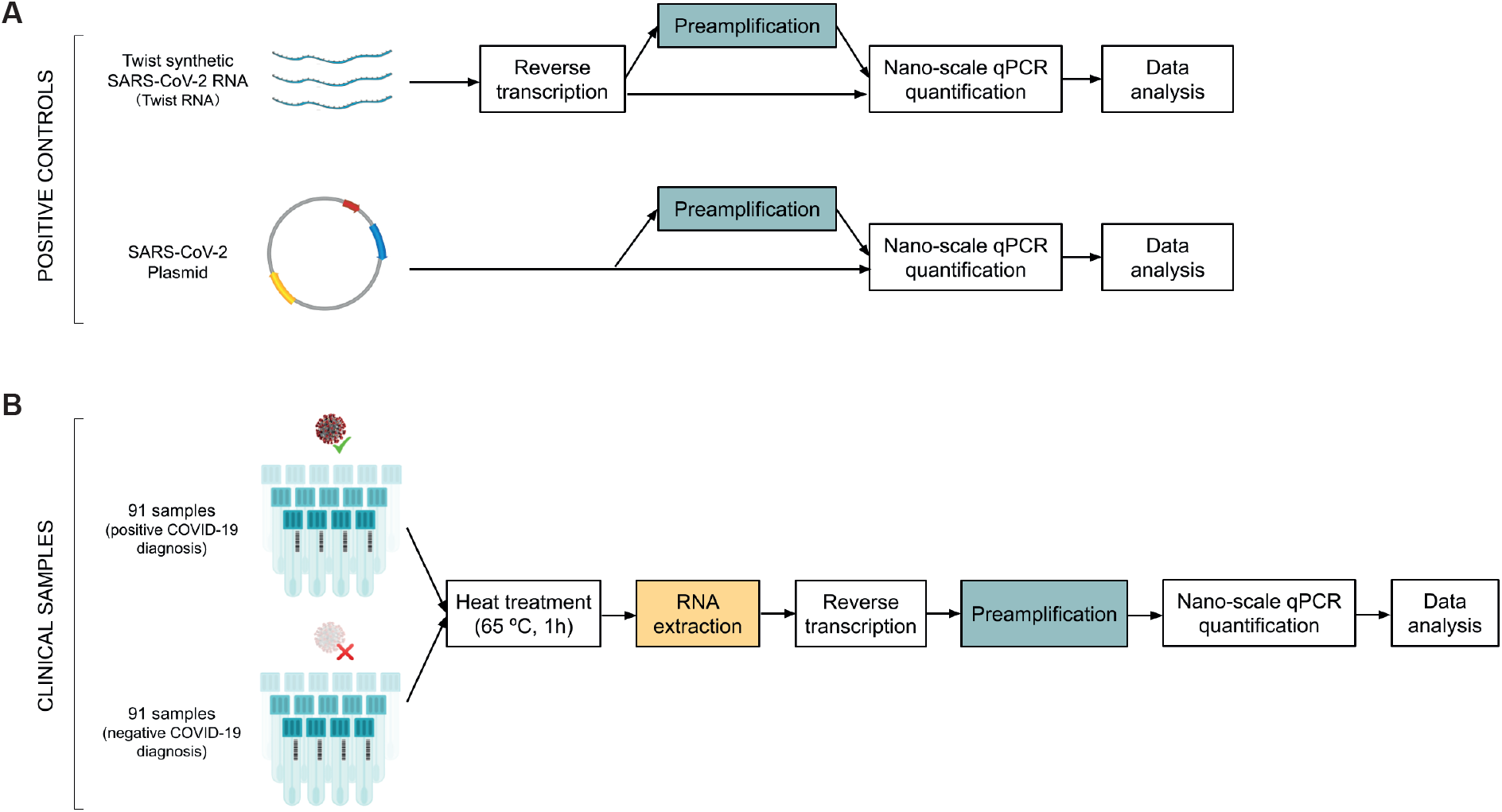
Schematic overview of the experimental design for SARS-CoV-2 RT-qPCR testing using the Fluidigm platform. (A) Workflow for method validation in positive controls (SARS-CoV-2 synthetic RNA and SARS-CoV-2 plasmids) with and without a preamplification step. (B) Workflow for clinically diagnosed SARS-CoV-2 samples (nasopharyngeal swabs) from an accredited diagnostic lab.

We first used 10-fold dilution series of the synthetic SARS-CoV-2 RNA (Twist RNA) and SARS-CoV-2 plasmid to determine the LoD for the N1 and N2 assays, with and without a preamplification step (Figure 2). With preamplification, the viral N gene was detectable in Twist RNA at 0.5 copies/μl (N1 assay) and 5 copies/μl (N2 assay), whereas without preamplification, we detected no viral material below a concentration of 5,000 copies/μl (Figure 2A). We observed similar results using the SARS-CoV-2 plasmid. (Figure 2B), which demonstrated that the preamplification step is essential for high detection sensitivity for SARS-CoV-2 in the range of 1 copy/ul. For this reason, we used a preamplification step in all subsequent experiments.

**Figure 2.**
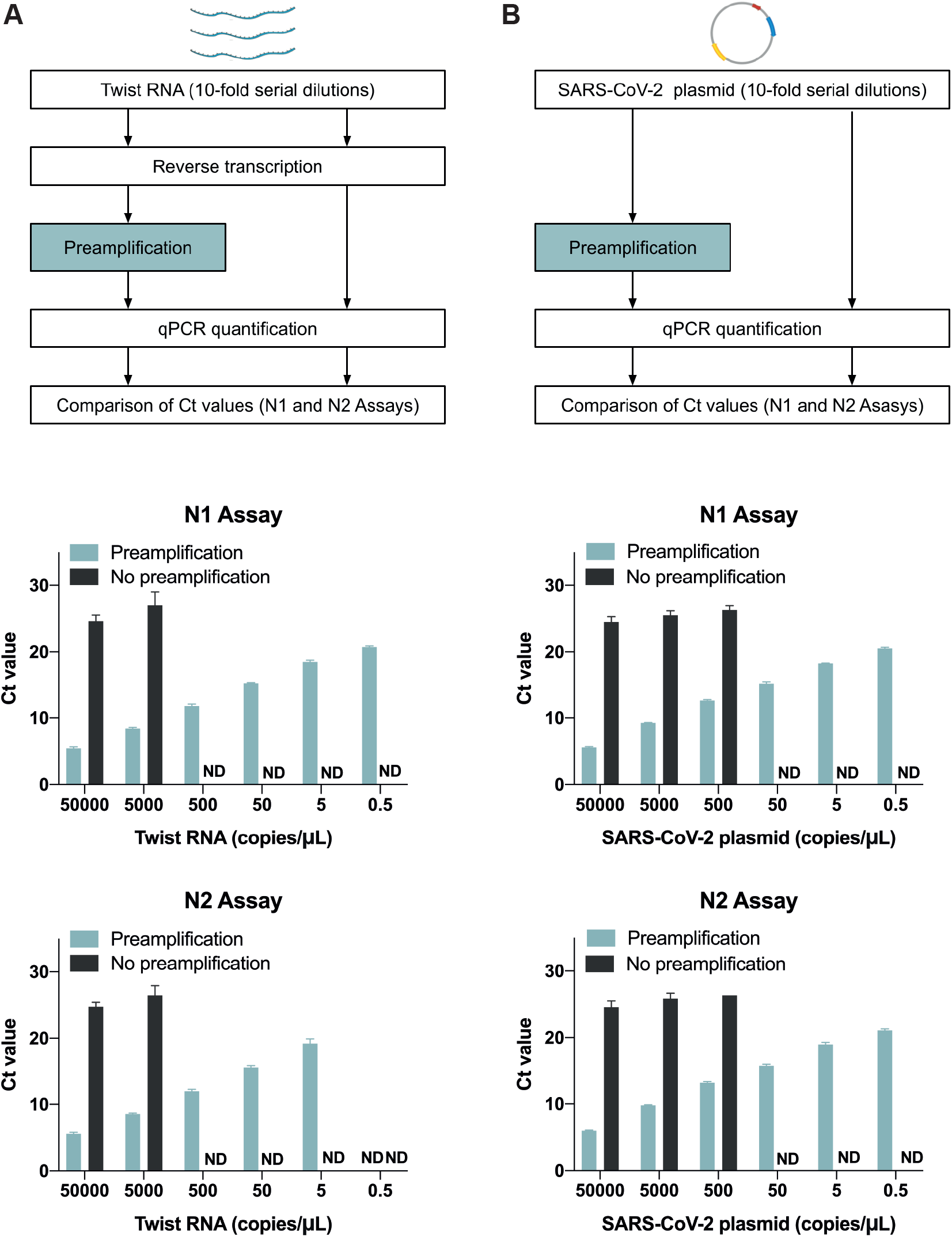
Preamplification leads to a lower limit of detection (LoD) in the Fluidigm method. (A, B) Workflow for 10-fold serially diluted SARS-CoV-2 positive controls, with and without preamplification: (A) synthetic RNAs (Twist RNA) and (B) SARS-CoV-2 plasmid. Barplots show the Ct values with and without the preamplification step for both the N1 and N2 assays at each concentration. Data show the Mean ± SD of valid Ct values from 18 replicates (2 replicates at each concentration, each with 9 technical replicates).

Since the LoD is dependent on the abundance of input material, we next examined the extent to which RNA extraction affects the quantity of input material. We extracted RNA from the Twist RNA dilution series and eluted in the same volume as the original samples (Figure S2). With or without extraction, we were able to detect 5 copies/μl using both the N1 and N2 assays, but, the Ct values for the extracted samples increased by 1-3 cycles at different dilutions. Most viral nucleic acid extraction kits recommend using carrier RNA to enhance recovery of viral RNA in samples where the quantity of material is low; however, carrier RNA might compete nonspecifically with the SARS-CoV-2 RNA in reverse transcription reactions. To explore this possibility, we added carrier RNA directly into the Twist RNA serial dilutions, and found that Ct values indeed increased by 0.9-2.7 cycles at most concentrations (Figure S2). We therefore conclude that the presence of carrier RNA, and not RNA extraction itself, can adversely affect the detection of viral material, possibly by interfering with the efficiency of reverse transcription.

To evaluate the sensitivity of our workflow in comparison with standard SARS-CoV-2 detection methods, we next analyzed 182 NP swab samples previously diagnosed by an accredited diagnostic laboratory as SARS-CoV-2 positive or negative (91 samples each). We heated the samples at 65 degrees for 1 hour to inactivate viral particles and analyzed them using our SARS-CoV-2 detection workflow (Figure 1B). Any samples with poor qPCR amplification curves and high variation in Ct values among replicates (SEM > 0.5) were flagged as inconsistent, which in our experience is usually due to technical issues such as the formation of air bubbles when loading the samples onto the chip. A total of 11 inconsistent samples (4 positive and 7 negative) were removed from subsequent analysis (Figure 3A). The remaining 171 high quality and valid samples were classified as either positive (either N1 or N2 detected) or negative (neither N1 nor N2 detected). Using our method, we confirmed 86 positive samples (Pos_Pos, 94.5%) among the 87 samples with a positive clinical diagnosis and found 1 negative (Pos_Neg, 1.1%) (Figure 3B). Among the 84 samples with a negative diagnosis, we detected 17 positives (Neg_Pos, 18.7%): in 9 of these samples, we obtained valid Ct values for all 18 replicates (both N1 and N2), and in the remaining 9, we detected the virus in 9/18 replicates (either all N1 or all N2). By performing a large number of replicates, our method can robustly detect SARS-CoV-2 and therefore identify samples as false negatives by the clinical diagnostic procedure.

**Figure 3.**
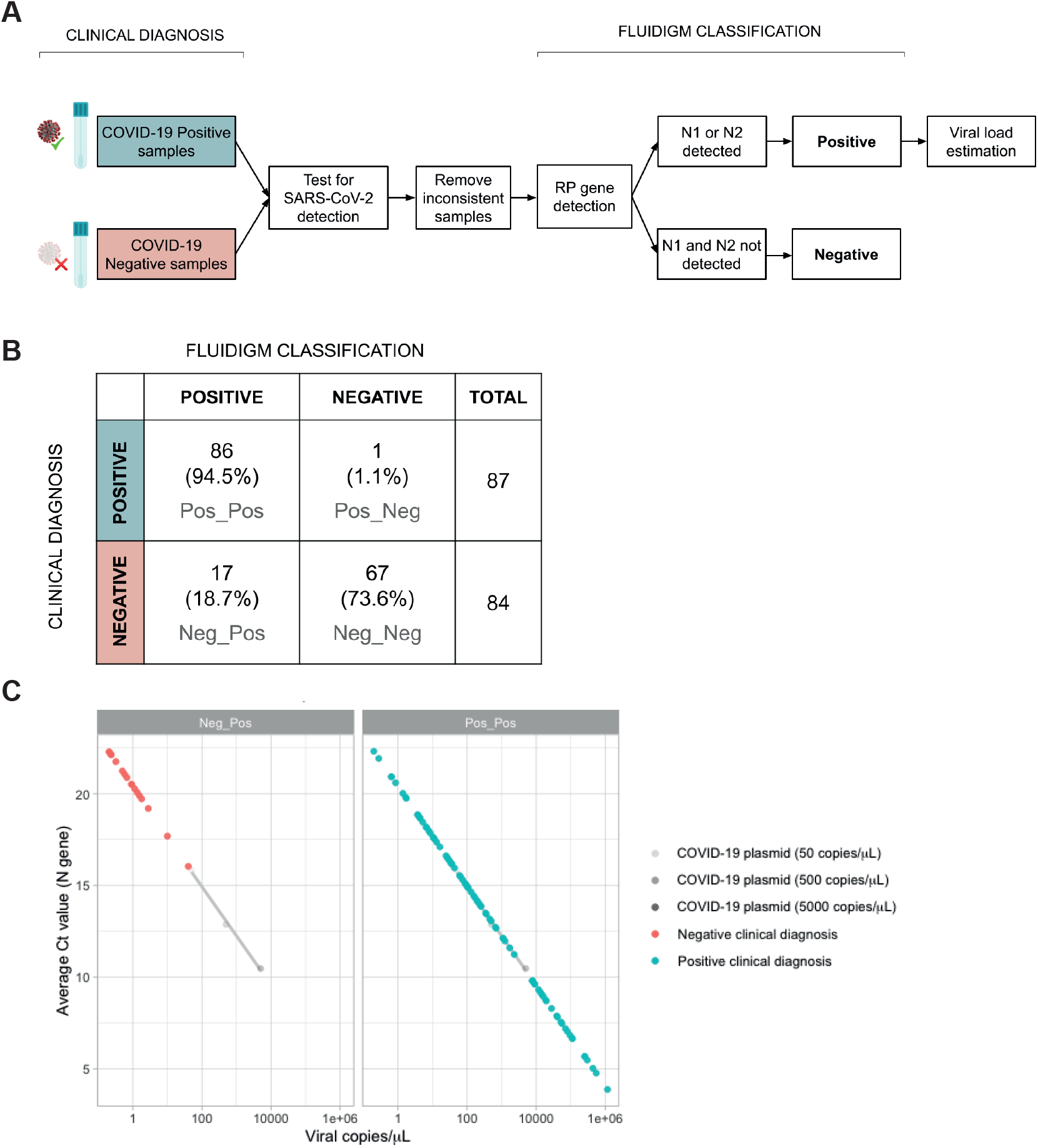
Three-step RT-qPCR assay with Fluidigm system shows higher sensitivity in detecting low viral-load samples. (A) Workflow for the re-analysis of clinical samples (SARS-CoV-2 positive and negative) from an accredited diagnostic lab. (B) Comparison between clinical diagnosis and Fluidigm classification. Category labels indicate clinical diagnosis followed by Fluidigm classification (e.g. Neg_Pos = negative diagnosis, positive Fluidigm result) (C) Viral load estimates for different categories of classified samples based on Fluidigm qPCR. Samples with a negative clinical diagnosis contained very low viral loads (<100 copies/μl). Viral titers of samples in Plate 1 and 2 were computed using the standard curve of plasmid controls from Plate 1.

Standard curves are commonly used to estimate viral loads in RT-qPCR reactions. Standard curves based on 100-fold serial dilutions of Twist RNA and SARS-CoV-2 plasmids ranging from 5-50,000 copies/μl both showed a nearly-perfect log-linear fit (R^2^ > 0.99) with little variation among technical replicates (SEM < 0.2) for both N assays (Figure S3). Having established technical precision using these curves, we used the SARS-CoV-2 plasmid standards to quantify viral copies in the clinical samples in each of the two PCR plates. Given that the N1 and N2 assays were highly concordant (R^2^=0.876, Kendall’s Tau correlation), and the difference in Ct values was within 1 cycle in over 90% of the samples (Figure S4A-B), we used an average Ct value for the N gene assays. To maximize consistency in viral load estimates among the samples, we then used the standard curve from plate 1 to quantify the viral loads in each clinical sample (Figure 3C). The Pos_Pos samples showed a wide range of estimated viral loads (0.2-1.17-10^6^ viral copies/μl) spanning five orders of magnitude. In contrast, the Neg_Pos samples exhibited a much narrower range of viral load estimates (0.2-40.25 viral copies/μl), corresponding to very low amounts of viral material in the NP swab sample (0.05-10.73 viral copies/ul). These results show that our nano-scale qPCR method can detect SARS-CoV-2 across a broad range of viral titers and can confidently detect relatively low viral loads that could otherwise be missed by standard SARS-CoV-2 detection methods used in diagnostic labs.

To evaluate the reproducibility of this method in samples with low viral loads, we re-analysed the 17 Neg_Pos samples after one and after two freeze cycles. After one freeze cycle, 11 samples remained positive, whereas after two freeze cycles, only 9 did (Figure 4A), suggesting that additional freeze cycles may compromise the power of our method to detect low viral loads, likely due to the degradation of viral RNA. Nevertheless, 8 of the 17 samples were reproducibly classified as positive in three independent experiments (Figure 4B). When comparing viral load estimates among the 17 Neg_Pos samples, we found that the 11 samples that retested positive after one freeze cycle had viral loads between 0.21-38.89 copies/μl, whereas the 5 samples that retested negative all had viral loads less than 1 copy/μl based on the original experiment (Figure 4C). This suggests that samples with extremely low viral titers close to the LoD could fail to be consistently detected, likely due to factors such as sample degradation or stochastic variation in the number of viral RNA molecules present in the small reaction volumes used for RT.

**Figure 4.**
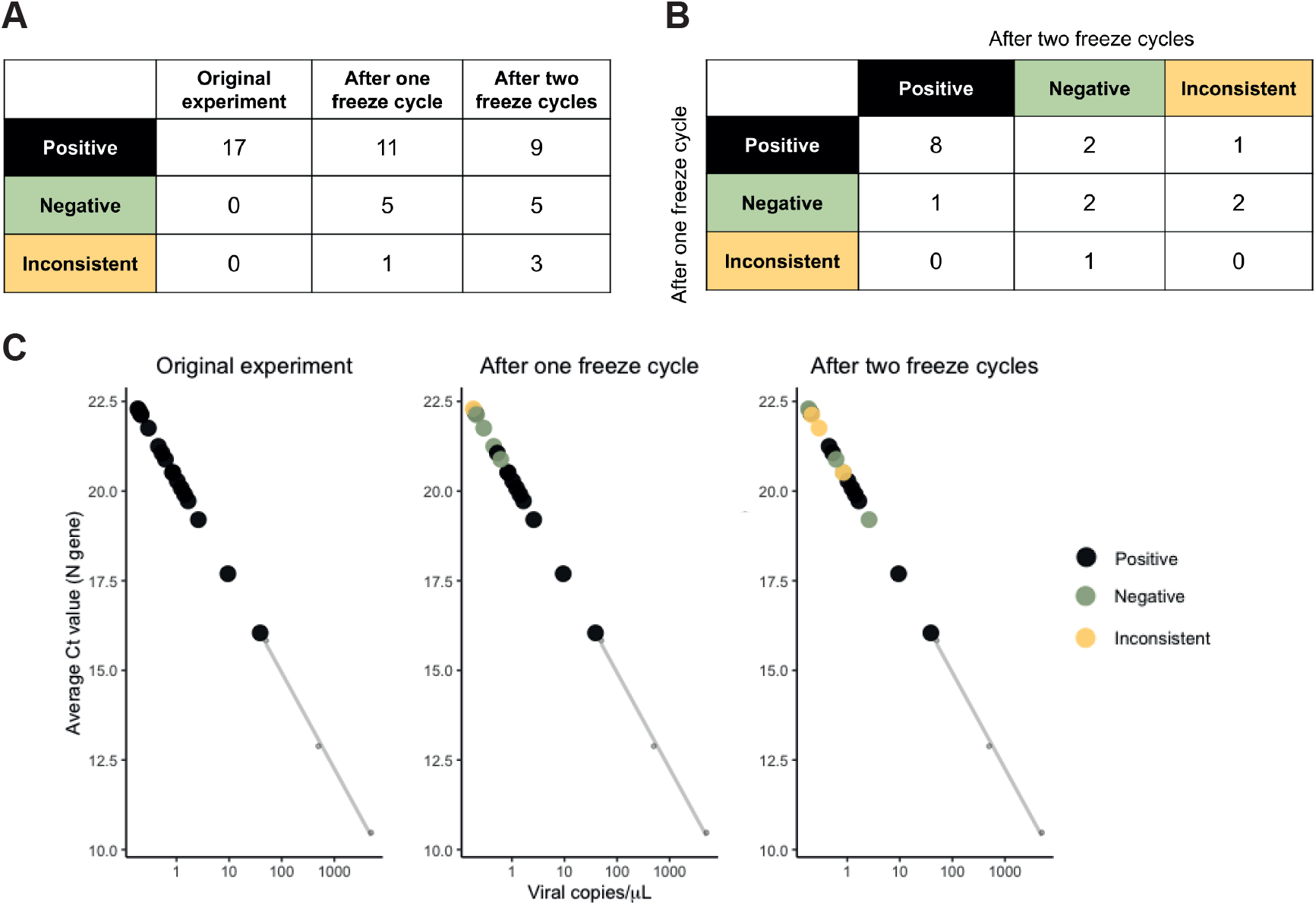
Reproducibility of results among the 17 Neg_Pos samples. (A) Summary of Fluidigm results across the original experiment and two replication experiments (after one freeze cycle and after two freeze cycles). (B) Cross-tabulation of Fluidigm results in the two replication experiments. (C) Viral load estimates for the 17 Neg_Pos samples based on the original experiment. Color-coding is based on the Fluidigm results of the 17 samples in the two replication experiments, as either positive (black), negative (green) or inconsistent (purple).

Lastly, we examined the relationship between the distributions of mean Ct values for the host RP gene assay and the N gene assay. If negative samples tended to show much higher Ct values for the RP assay than the positive samples, this would indicate that the negative result was likely due to inadequate sampling of human tissues in the swab. Instead, we observed no significant differences in the Ct values of RP assay between positive and negative samples (t-test, p-value = 0.08) (Figure 5A) and found minimal correlation between the mean Ct values of the RP and the N gene assays, with only 2.8% of the variation in the N gene assays attributable to detection of the RP gene (R^2^=0.028, Kendall’s Tau correlation) (Figure 5B). We thus conclude that the high Ct values for the N assays in the Neg_Pos samples were not due to inadequate sampling, but instead accurately reflected low viral loads in these samples.

**Figure 5.**
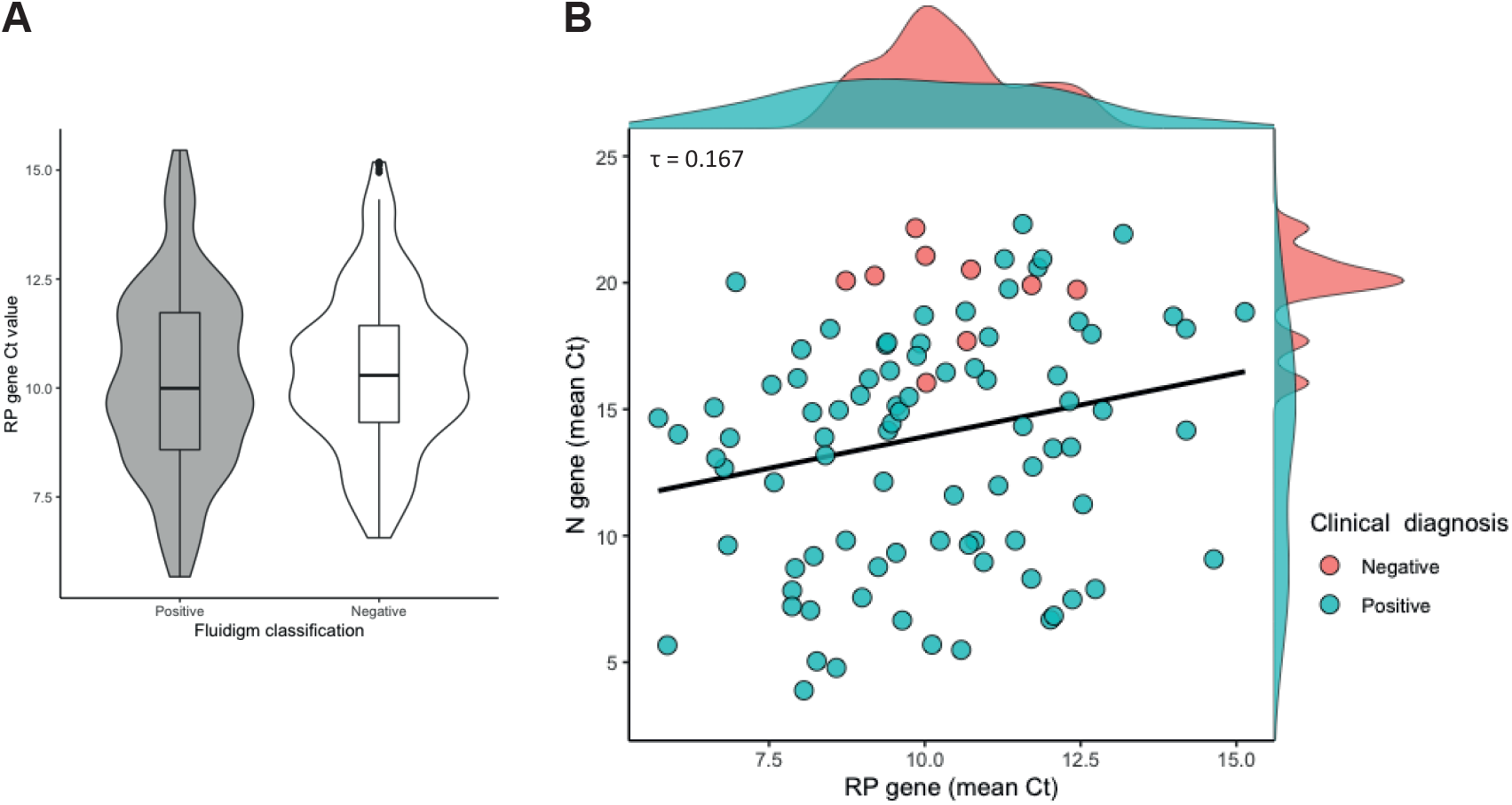
Low estimated viral titers are not due to poor quality samples. (A) Violin plots of RP gene Ct values across samples with positive and negative Fluidigm results. There is no significant difference between the two groups (Welch’s t-test, p-value = 0.08). (B) Scatter and density plots of N gene vs. RP gene Ct values from Fluidigm qPCR for samples with negative or positive clinical diagnostic results. Ct values for viral N and host RP genes are not strongly correlated overall (τ = 0.167, p-value = 0.017).

## Discussion

In this study, we implement and validate a three-step approach for SARS-CoV-2 detection utilizing RT of SARS-CoV-2 viral RNA, cDNA preamplification and nano-scale qPCR. Using serial dilutions of positive controls, we demonstrate a 1,000-fold improvement in detection sensitivity of the Fluidigm system when adding the preamplification step, and we show that nano-scale qPCR can be used to quantify viral copies in clinical samples with high confidence. Our data suggests that the LoD of this method (with preamplification) is less than 1 copy/μl: we obtained an LoD of 0.5 copies/μl for Twist RNA and SARS-CoV-2 plasmid, and detected the virus down to 0.2 copies/μl in experiments using clinical NP swab samples. Based on this analysis, our method seems to be more sensitive than standard RT-PCRs, which have reported a LoD ranging between 5.6 and 100 copies/μl [23-25], and that its performance is comparable to other highly sensitive methods such as the CDC 2019-nCoV RT-PCR Diagnostic Panel with QIAGEN QIAmp DSP Viral RNA Mini Kit and the QIAGEN EZ1 DSP [3] (1 copy/μl), ddPCR (0.1 copies/μl) [8], and RT-LAMP (1 copy/μl) [26].

Crucially, we demonstrate the power of this method to reduce the false negative rate of SARS-CoV-2 clinical diagnostic tests: we detected SARS-CoV-2 in 17 samples diagnosed as negative by an accredited diagnostic lab with high confidence, based on a large number of technical replicates (9 for N1, 9 for N2, 6 for RP) and a conservative threshold for Ct value consistency among sample replicates (SEM < 0.5). This is especially important considering that the viral loads of these Neg_Pos samples (0.2-40.25 viral copies/μl) were close to the LoD of standard SARS-CoV-2 tests and are thus more likely to return false negative results using standard RT-PCR methods. This three-step microfluidics RT-qPCR method, which includes a preamplification step, nano-scale reactions and a large number of replicates can reliably detect samples with low viral load and reduce the false negative rate compared to standard RT-PCR assays.

Beyond its ultra-sensitive detection of SARS-CoV-2, this microfluidic platform is cost-effective strategy with several advantages: a nanoliter volume per reaction (lower reagent consumption per assay), a parallelized assay system (increased throughput), amenability to automation (increased precision), capacity to run a large number of replicates per sample (increased confidence in test results) and the capacity to simultaneously test for multiple pathogens (broader diagnostic utility). Because of this, the method has great potential for economies of scale, including assay multiplexing (to detect additional pathogens) and sample pooling (to increase throughput), which would further reduce per-test costs. These advantages warrant serious consideration of this three-step nano-scale assay system, especially for active screening programs which aim at the early detection of SARS-CoV-2 in asymptomatic, pre-asymptomatic or mildly symptomatic individuals who likely carry low viral loads.

## Data Availability

All data referred to in the manuscript (q-RT-PCR data) is available on request.

## Acknowledgements

This report is part of NYUAD’s COVID Response Project. We thank members of NYU Abu Dhabi’s (NYUAD) COVID-19 steering committee for spearheading and facilitating this project, including Drs. Ayaz Virji and Sehamuddin Galadari. We thank members of Proficiency Healthcare Diagnostics laboratory for sample collection and processing. We thank Michael Davis (Director of Laboratory Operations, NYUAD), Raza Rowshan (Director of Core Technology Platforms, NYUAD), Nizar Drou (Lead Developer, NYUAD Bioinformatics Core), Nada Messaikeh (Vice Provost for Research Administration, NYUAD), and Enas Qudeimat (Director of Operations, NYUAD-CGSB) for helping to make this work possible. We thank the Institutional Review Boards of the Abu Dhabi Department of Health and NYUAD and the NYUAD Institutional Biosafety Committee for expediting review of this project and NYUAD Environmental Health and Safety for their support.

## Author Contributions

K.C.G., Y. I., and F.P. conceived and supervised the project. X.X., Z.A., M.M.D., M.A., M.K., Y.M. and F.A.J. performed the experiments of positive controls and clinical sample processing. K.C.G. and N.R. designed the database; N.R. implemented and C.A.J. contributed to the infrastructure for data curation and management. Z.V. and M.Z. provided clinical samples for this study. T.G. and X.X. performed data analysis. X.X., T.G., K.C.G., Y.I., Z.A. and M.M.D. contributed to data interpretation and manuscript preparation. F.P., K.C.G., Y. I. and R.A. acquired funding and approvals for this study.

